# COVID-19 in Wuhan: Immediate Psychological Impact on 5062 Health Workers

**DOI:** 10.1101/2020.02.20.20025338

**Authors:** Zhou Zhu, Shabei Xu, Hui Wang, Zheng Liu, Jianhong Wu, Guo Li, Jinfeng Miao, Chenyan Zhang, Yuan Yang, Wenzhe Sun, Suiqiang Zhu, Yebin Fan, Junbo Hu, Jihong Liu, Wei Wang

**Affiliations:** Department of Neurology, Tongji Hospital, Tongji Medical College, Huazhong University of Science and Technology, No. 1095 Jiefang Avenue, Qiaok’ou District, Wuhan 430030, Hubei, China, or at; Department of Urology, Tongji Hospital, Tongji Medical College, Huazhong University of Science and Technology, No. 1095 Jiefang Avenue, Qiaokou District, Wuhan 430030, Hubei, China, or at; Department of Gastrointestinal Surgery, Tongji Hospital, Tongji Medical College, Huazhong University of Science and Technology, No. 1095 Jiefang Avenue, Qiaokou District, Wuhan 430030, Hubei, China, or at; Department of Psychiatry, Wuhan, Hubei, China; Department of Neurology, Wuhan, Hubei, China; Nursing Department, Wuhan, Hubei, China; Department of Otolaryngology–Head and Neck Surgery, Wuhan, Hubei, China; Department of Gastrointestinal Surgery, Wuhan, Hubei, China; Department of Urology, Wuhan, Hubei, China; Tongji Hospital, Tongji Medical College, Huazhong University of Science and Technology, and the School of Computer Science and Technology, Huazhong University of Science and Technology, Wuhan, Hubei, China

## Abstract

**BACKGROUND:** The outbreak of COVID-19 has laid unprecedented psychological stress on health workers (HWs). We aimed to assess the immediate psychological impact on HWs at Tongji Hospital in Wuhan, China.

**METHODS:** We conducted a single-center, cross-sectional survey of HWs via online questionnaires between February 8th and 10th, 2020. We evaluated stress, depression and anxiety by Impact of Event Scale-Revised (IES-R), Patient Health Questionnaire-9 (PHQ-9), and Generalized Anxiety Disorder 7-item (GAD-7), respectively. We also designed a questionnaire to assess the effect of psychological protective measures taken by Tongji Hospital. Multivariate logistic regression was used to identify predictors of acute stress, depression, and anxiety.

**RESULTS:** We received 5062 completed questionnaires (response rate, 77.1 percent). 1509 (29.8 percent), 681 (13.5 percent) and 1218 (24.1 percent) HWs reported stress, depression and anxiety symptoms. Women (hazard ratio[HR], 1.31; P=0.032), years of working> 10 years (HR, 2.02; P<0.001), concomitant chronic diseases (HR, 1.51; P<0.001), history of mental disorders (HR, 3.27; P<0.001), and family members or relatives confirmed or suspected (HR, 1.23; P=0.030) were risk factors for stress, whereas care provided by hospital and department administrators(odds ratio [OR], 0.76; P=0.024) and full coverage of all departments with protective measures (OR, 0.69; P=0.004) were protective factors.

**CONCLUSIONS:** Women and those who have more than 10 years of working, concomitant chronic diseases, history of mental disorders, and family members or relatives confirmed or suspected are susceptible to stress, depression and anxiety among HWs during the COVID-19 pandemic. Psychological protective measures implemented by the hospital could be helpful.

## BACKGROUND

At the end of 2019, the COVID-19 emerged in Wuhan City, Hubei Province, China. The rapid escalation of COVID-19 epidemic has resulted in a World Health Organization (WHO)-declared public health emergency of international concern. The global total number of COVID-19 cases has been several times that of SARS, and the death toll has also exceeded that of SARS^1^. As the source area of COVID-19 epidemic, Wuhan has the majority of confirmed cases and deaths worldwide. Nowadays, information is spreading more rapidly and extensively than it was in 2003 when SARS broke out, which might exacerbate public fear, panic, and distress. Frontline health workers (HWs) were saving lives while encountering an increasing workload and risk of infection. In the early stage of COVID-19 epidemic, it was reported that infected HWs accounted for 29 percent of all hospitalized COVID-19 patients^2^. Also, quarantined frontline HWs might be facing potential social isolation, and unquarantined HWs experiencing social discrimination. Therefore, they are susceptible to complex emotional reactions and psychological distress^3^. Furthermore, the mental health problems of HWs would impair their attention, cognitive functioning, and clinical decision-making^4,5^, consequently increase the occurrence of medical errors and incidents, and ultimately put patients at risk. It was also well known that acute stress in disasters could have a lasting effect on the overall wellbeing^6-8^. Hence, the mental health problems of HWs in COVID-19 epidemic have become an urgent public health concern.

Tongji Hospital, one of the biggest tertiary hospitals in Wuhan, has been designated by the government as “the specific hospital for the treatment of severe patients with COVID-19 in Wuhan”. Since the outbreak of COVID-19, Tongji hospital has opened 2,000 beds specifically for severe COVID-19 patients, and continually transferred HWs from all departments to the frontline of COVID-19, with a total of over 3000 HWs. The management of Tongji Hospital was initially alert to the mental health problems that HWs might encounter, and therefore implemented a series of psychological protective measures, including organizing WeChat Balint group, providing hospital-based, department-based, and ward-based care, full coverage of all departments with protective measures for nosocomial infection, reasonable work shift arrangement, and sufficient logistical support and comfortable accommodations for HWs.

On Jan 26, 2020, China’s National Health Commission (NHC) issued Guidelines for Emergency Psychological Crisis Interventions during the COVID-19 Epidemic^*9*^. To date, research on the immediate psychological impact of COVID-19 on HWs is still lacking. We aimed to evaluate the immediate psychological impact on HWs at Tongji Hospital, determine the predictors of acute stress, depression, and anxiety, assess the effectiveness of psychological protective measures, and develop effective, easy-to-use clinical screening tools to identify high-risk individuals of acute stress, depression, and anxiety among HWs.

## METHODS

### STUDY DESIGN AND PARTICIPANTS

The study was a single-center, cross-sectional survey, covering doctors, nurses and clinical technicians in all clinical departments of Tongji Hospital. The study was conducted between February 8th and 10th, 2020, two weeks after the authority in Wuhan suspended all public transport on January 23. The questionnaire consisted of five parts: online informed consent, baseline sociodemographic information, perceptions of threat of COVID-19, psychological protective measures and rating scales including Impact of Event Scale-Revised Questionnaires (IES-R), Patient Health Questionnaire-9 (PHQ-9), and Generalized Anxiety Disorder 7-item (GAD-7).

Data were collected through anonymous online questionnaires which were distributed to all HWs via WeChat. Only one response per person to the questionnaire was permitted. The senior investigators performed quality control by checking the collected questionnaires daily. Two researchers entered data into the database double-blindly using Epidata.3.0 to guarantee accuracy. The study was approved by the institutional ethics board of Tongji Hospital, Tongji Medical College of Huazhong University of Science and Technology (ID: TJ-C20200129). The data analyses were done on unidentified datasets.

### MEASURES

The sociodemographic information was collected. The frontline HWs are those directly providing services to confirmed or suspected COVID-19 patients. Exercise habits are defined as meeting the WHO physical activity recommendations for adults aged 18-64 years old^10^.

We provide five items to assess the perceptions of threat of COVID-19: (1) Do you feel that you have a history of exposure to COVID-19? (2) Have you ever thought of resigning because of COVID-19 outbreak? (3) Have you worried about the life-threatening once infected? (4) Do you feel that families and friends have avoided contact with you because of your work? (5) Have you worried about yourself or your family members being infected by COVID-19?

The psychological protective measures included the following 5 items: (1) Have you joined Tongji WeChat Balint Group? (2) Have you received care provided by hospital and department administrators? (3) Are you satisfied with full coverage of all departments with protective measures for nosocomial infection? (4) Are you satisfied with your work shift arrangement?(5) Are you satisfied with the logistical support and comfortable accommodations provided by Tongji Hospital?

### OUTCOMES

PHQ-9 Scale was used to measure the depression symptoms. A cutoff of ≥10 has been recommended for diagnosis of major depression, which provides adequate sensitivity (88.0 percent) and specificity (88.0 percent)^11^. GAD-7 Scale was used to identify anxiety disorders. A cutoff score ≥8 is recommended to identify clinically important anxiety symptoms, with adequate specificity (82.0 percent) and sensitivity (77.0 percent)^12^. IES-R is a 22-item self-reported measure applied to assess subjective stress caused by traumatic event, describing the three symptoms including avoidance, intrusion, and hyperarousal. We defined the psychological stress when the IES-R score >33 points^13^. The selected 3 questionnaires had good internal consistency with Cronbach’s α coefficients of more than 0.80.

### STATISTICAL ANALYSIS

Continuous variables were divided as categorical variables firstly and all variables were shown as the counts and percentages. Variables with P values<0.05 in univariate analyses were subjected to multivariate logistic regression analysis with a stepwise backwards elimination procedure. Statistical analyses to identify influencing factors were performed using SPSS 22.0 (Statistical Package for the Social Sciences) for Windows (SPSS, Chicago, IL). The Depression, Anxiety and Psychological Stress nomograms were formulated based on the results of multivariate logistic regression analysis using the R packages “rms”, “Hmisc” and “ggplot2” in R 3.5.2 (http://www.r-project.org/). The performances of the nomograms were measured by Concordance statistics (C-statistics) and assessed by calibration curves. Bootstraps with 1000 resamples were applied to these activities. The higher C-statistics indicate better ability to distinguish HWs with different risks of three outcomes. We considered the risk screening models as a useful clinical tool particularly when the C-statistic is higher than 0.70^14^. The calibration curves were used to compare the observed probability with the predicted probability. Dots on the calibration plot would be close to a 45° diagonal line if the model calibration is correct.

## RESULTS

6568 individuals were surveyed and 5281 individuals completed the online questionnaire. 5062 HWs were included in the final analysis after excluding the 219 questionnaires with wrong information (response rate, 77.1 percent). Most subjects are in the age intervals of 19-29 (40.1 percent) and 30-49 years old (56.4 percent). The females accounted for 85.0 percent of the total respondents. Baseline characteristics for the total sample (n=5062), those depressive HWs (n=681, 13.5 percent) vs. non-depressive ones (n=4381), anxious HWs (n=1218, 24.1 percent) vs. non-anxious ones (n=3844), psychological stressed (n=1509, 29.8 percent) vs. non-psychological stressed (n = 3553) at baseline were presented in Table 1. We also summarized the perceptions of the threat of COVID-19 among HWs and psychological protective measures, as shown in Table 2. The baseline characteristics of subgroups were presented in Supplementary Appendix Tables S1-2, S4-5 and S7-8.

**Table 1.** Sociodemographic characteristics of Wuhan HWs dealing with the COVID-19

| Characteristics | Total<br>(N=5062) | Depression<br>(N= 681) | Non-depression<br>(N=4381) | P | Anxiety<br>(N=1218) | Non-anxiety<br>(N=3844) | P | Psychological stress<br>(N=1509) | Non-psychological<br>stress<br>(N=3553) | P |
| --- | --- | --- | --- | --- | --- | --- | --- | --- | --- | --- |
| Age |  |  |  | 0.006 |  |  | < 0.001 |  |  | < 0.001 |
| 19~29 years | 2030 (40.1%) | 235 (34.5%) | 1795 (41.0%) |  | 385 (31.6%) | 1645 (42.8%) |  | 518 (34.3%) | 1512 (42.6%) |  |
| 30~49 years | 2854 (56.4%) | 421 (61.8%) | 2433 (55.5%) |  | 794 (65.2%) | 2060 (53.6%) |  | 944 (62.6%) | 1910 (53.8%) |  |
| > 49 years | 178 (3.5%) | 25 (3.7%) | 153 (3.5%) |  | 39 (3.2%) | 139 (3.6%) |  | 47 (3.1%) | 131 (3.7%) |  |
| Women | 4304 (85.0%) | 610 (89.6%) | 3771 (86.1%) | < 0.001 | 1081 (88.8%) | 3223 (83.8%) | < 0.001 | 1356 (89.9%) | 2948 (83.0%) | < 0.001 |
| Marital status |  |  |  | 0.044 |  |  | < 0.001 |  |  | < 0.001 |
| Married | 3356 (66.3%) | 479 (70.3%) | 2877 (65.7%) |  | 895 (73.5%) | 2461 (64.0%) |  | 1072 (71.0%) | 2284 (64.3%) |  |
| Unmarried | 1576 (31.1%) | 184 (27.0%) | 1392 (31.8%) |  | 287 (23.6%) | 1289 (33.5%) |  | 394 (26.1%) | 1182 (33.3%) |  |
| Divorced/Widowed/ Separated | 130 (2.6%) | 18 (2.6%) | 112 (2.6%) |  | 36 (2.0%) | 94 (2.5%) |  | 43 (2.9%) | 87 (2.5%) |  |
| Master degree or higher level | 1235 (24.4%) | 150 (22.0%) | 1085 (24.8%) | 0.121 | 293 (24.1%) | 942 (24.5%) | 0.75 | 325 (21.5%) | 910 (25.6%) | 0.002 |
| Occupation |  |  |  | 0.032 |  |  | 0.003 |  |  | < 0.001 |
| Doctor | 1004 (19.8%) | 114 (16.7%) | 890 (20.3%) |  | 232 (19.1%) | 772 (20.1%) |  | 243 (16.1%) | 761 (21.4%) |  |
| Nurse | 3417 (67.5%) | 489 (71.8%) | 2928 (66.8%) |  | 863 (70.9%) | 2554 (66.4%) |  | 1130 (74.9%) | 2287 (64.4%) |  |
| Medical technician | 641 (12.7%) | 78 (11.5%) | 563 (12.9%) |  | 123 (10.3%) | 518 (13.48%) |  | 136 (9.0%) | 505 (14.2%) |  |
| Professional title level |  |  |  | < 0.001 |  |  | < 0.001 |  |  | < 0.001 |
| Junior | 3010 (59.5%) | 359 (52.7%) | 2651 (60.5%) |  | 634 (52.1%) | 2376 (61.81%) |  | 843 (55.9%) | 2167 (61.0%) |  |
| Intermediate | 1664 (32.9%) | 270 (39.7%) | 1394 (31.8%) |  | 478 (39.2%) | 1186 (30.85%) |  | 563 (37.3%) | 1101 (31.0%) |  |
| Senior | 388 (7.7%) | 52 (7.6%) | 336 (7.7%) |  | 106 (8.7%) | 282 (7.3%) |  | 103 (6.8%) | 285 (8.0%) |  |
| Years of working |  |  |  | < 0.001 |  |  | < 0.001 |  |  | < 0.001 |
| < 2 years | 505 (10.0%) | 45 (6.6%) | 460 (10.5%) |  | 77 (6.3%) | 428 (11.1%) |  | 92 (6.1%) | 413 (11.6%) |  |
| 2~5 years | 1724 (34.1%) | 201 (29.5%) | 1523 (34.8%) |  | 338 (27.8%) | 1386 (36.1%) |  | 454 (30.1%) | 1270 (35.7%) |  |
| 6~10 years | 1376 (27.2%) | 180 (26.4%) | 1196 (27.3%) |  | 354 (29.1%) | 1022 (26.6%) |  | 455 (30.2%) | 921 (25.9%) |  |
| > 10 years | 1457 (28.8%) | 255 (37.4%) | 1202 (27.4%) |  | 449 (36.9%) | 1008 (26.2%) |  | 508 (33.7%) | 949 (26.7%) |  |
| Frontline at present | 1736 (34.3%) | 240 (35.2%) | 1496 (34.2%) | 0.576 | 416 (34.2%) | 1320 (34.3%) | 0.906 | 582 (38.6%) | 1154 (32.5%) | < 0.001 |
| Current work place |  |  |  | 0.478 |  |  | 0.642 |  |  | < 0.001 |
| Non-isolation Ward | 3240 (64.0%) | 426 (62.6%) | 2814 (64.2%) |  | 767 (63.0%) | 2473 (64.3%) |  | 907 (60.1%) | 2333 (65.7%) |  |
| Isolation Ward | 1607 (31.8%) | 229 (33.6%) | 1378 (31.5%) |  | 400 (32.8%) | 1207 (31.4%) |  | 553 (36.7%) | 1054 (29.7%) |  |
| Off work or in isolation | 215 (4.3%) | 26 (3.8%) | 189 (4.3%) |  | 51 (4.2%) | 164 (4.3%) |  | 49 (3.2%) | 166 (4.7%) |  |
| Department |  |  |  | 0.852 |  |  | 0.313 |  |  | < 0.001 |
| Fever clinic, ED or isolation wards | 1741 (34.3%) | 239 (35.1%) | 1502 (34.3%) |  | 412 (33.8%) | 1329 (34.6%) |  | 582 (38.6%) | 1159 (32.6%) |  |
| Non-isolation wards | 2614 (51.6%) | 351 (51.5%) | 2263 (51.7%) |  | 649 (53.3%) | 1965 (51.1%) |  | 749 (49.6%) | 1865 (52.5%) |  |
| Other department | 707 (14.0%) | 91 (13.4%) | 616 (14.1%) |  | 157 (12.9%) | 550 (14.3%) |  | 178 (11.8%) | 529 (14.90%) |  |
| Annual household income |  |  |  | 0.65 |  |  | 0.001 |  |  | 0.046 |
| 30~100 thousand Yuan | 850 (16.8%) | 104 (15.3%) | 746 (17.0%) |  | 170 (14.0%) | 680 (17.7%) |  | 219 (14.5%) | 631 (17.8%) |  |
| 100~200 thousand Yuan | 2276 (45.0%) | 314 (46.1%) | 1962 (44.8%) |  | 529 (43.4%) | 1747 (45.5%) |  | 698 (46.3%) | 1578 (44.4%) |  |
| 200~300 thousand Yuan | 1237 (24.4%) | 172 (25.3%) | 1065 (24.3%) |  | 328 (26.9%) | 909 (23.7%) |  | 378 (25.1%) | 859 (24.2%) |  |
| > 300 thousand Yuan | 699 (13.8%) | 91 (13.4%) | 608 (13.9%) |  | 191 (15.7%) | 508 (13.2%) |  | 214 (14.2%) | 485 (13.7%) |  |
| Living at home | 3758 (74.2%) | 504 (74.0%) | 3254 (74.3%) | 0.892 | 908 (74.6%) | 2850 (74.1%) | 0.763 | 1073 (71.1%) | 2685 (75.6%) | 0.001 |
| Living with family member | 2514 (49.7%) | 346 (50.8%) | 2168 (49.5%) | 0.521 | 646 (53.0%) | 1868 (48.6%) | 0.007 | 721(47.8%) | 1793 (50.5%) | 0.081 |
| Past medical history |  |  |  | < 0.001 |  |  | < 0.001 |  |  | < 0.001 |
| In good health | 4158 (82.1%) | 488 (71.7%) | 3670 (83.8%) |  | 899 (73.8%) | 3259 (84.8%) |  | 1132 (75.0%) | 3026 (85.2%) |  |
| Have chronic non-communicable diseases | 848 (16.8%) | 161 (23.6%) | 687 (15.7%) |  | 281 (23.1%) | 567 (14.8%) |  | 340 (22.5%) | 508 (14.3%) |  |
| Have history of mental disorders | 56 (1.1%) | 32 (4.7%) | 24 (0.6%) |  | 38 (3.1%) | 18 (0.5%) |  | 37 (2.5%) | 19 (0.5%) |  |
| Smoking | 148 (2.9%) | 17 (2.5%) | 131(3.0%) | 0.477 | 25 (2.1%) | 123 (3.2%) | 0.038 | 31(2.1%) | 117 (3.3%) | 0.017 |
| Drinking | 426 (8.4%) | 73 (10.7%) | 353 (8.1%) | 0.02 | 113 (9.3%) | 313 (8.1%) | 0.214 | 126 (8.3%) | 300 (8.4%) | 0.913 |
| Exercise habit | 857 (16.9%) | 93 (13.7%) | 764 (17.4%) | 0.014 | 151 (12.4%) | 613 (16.0%) | < 0.001 | 209 (13.9%) | 648 (18.2%) | < 0.001 |
| Parent status |  |  |  | < 0.001 |  |  | < 0.001 |  |  | < 0.001 |
| No child | 1950 (38.5%) | 224 (32.9%) | 1726 (39.4%) |  | 366 (30.1%) | 1584 (41.2%) |  | 479 (31.7%) | 1471 (41.4%) |  |
| One child | 2444 (48.3%) | 340 (49.9%) | 2104 (48.0%) |  | 640 (52.6%) | 1804 (46.9%) |  | 775 (51.4%) | 1669 (47.0%) |  |
| Two or more children | 668 (13.2%) | 117 (17.2%) | 551 (12.6%) |  | 212 (17.4%) | 456 (11.9%) |  | 255 (16.9%) | 413 (11.6%) |  |
| Families or relatives suspected or confirmed | 763 (15.1%) | 146 (21.4%) | 617 (14.1%) | < 0.001 | 258 (21.2 %) | 505 (13.1%) | < 0.001 | 291 (19.3%) | 472 (13.3%) | < 0.001 |
| Suspected or confirmed COVID-19 | 158 (3.1%) | 41 (6.0%) | 117 (2.7%) | < 0.001 | 56 (4.6%) | 102 (2.7%) | 0.001 | 59 (3.9%) | 99 (2.8%) | 0.035 |

**Table 2.**
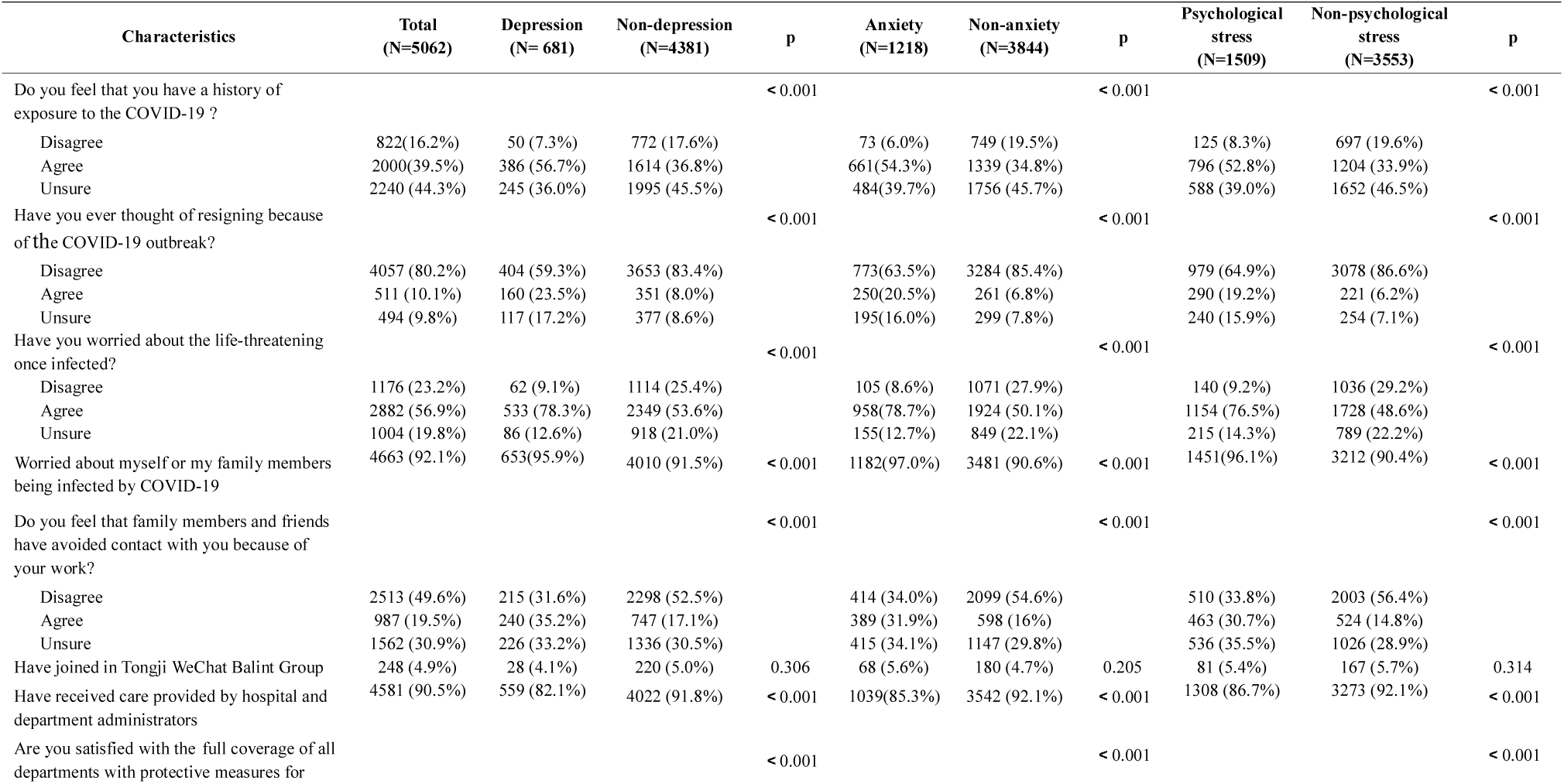
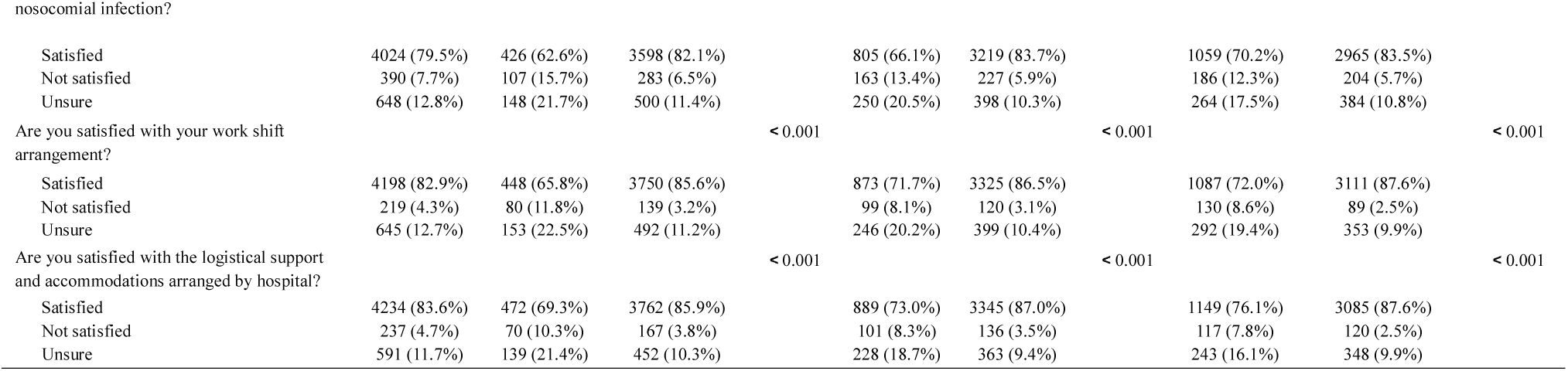
Perceptions of threat of the COVID-19 and effects of psychological protective measures among Wuhan HWs dealing with the COVID-19

Four sociodemographic variables presented as the common risk factors of depression, anxiety and psychological stress symptoms in HWs (all P values<0.05, Table 3). Take stress outcome as example, the details of risk factors were as following: women (hazard ratio [HR], 1.31, P=0.03), years of working> 10 years (HR, 2.02, P<0.001), concomitant chronic diseases(HR, 1.51; P<0.001), history of mental disorders (HR, 3.27; P<0.001), and family members or relatives confirmed or suspected (HR, 1.23; P=0.03).

**Table 3.**
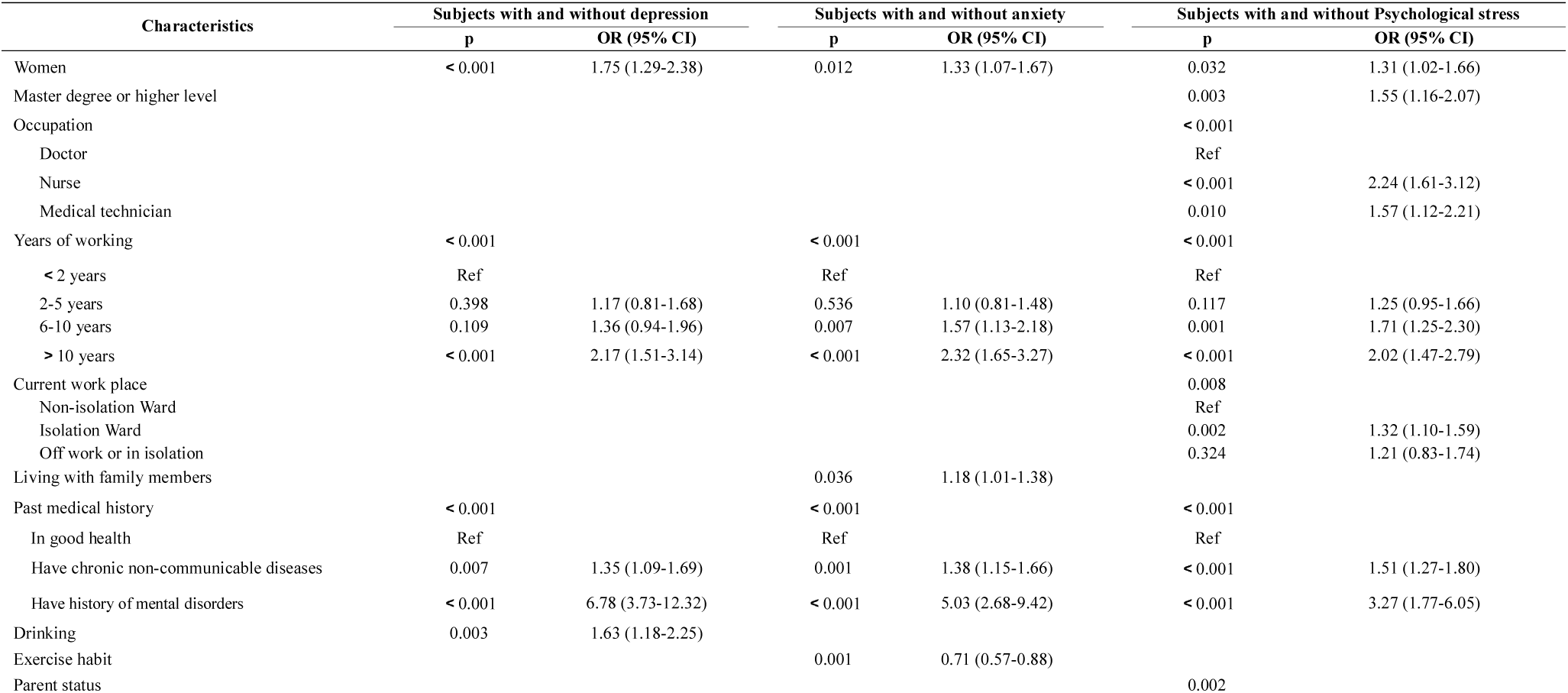
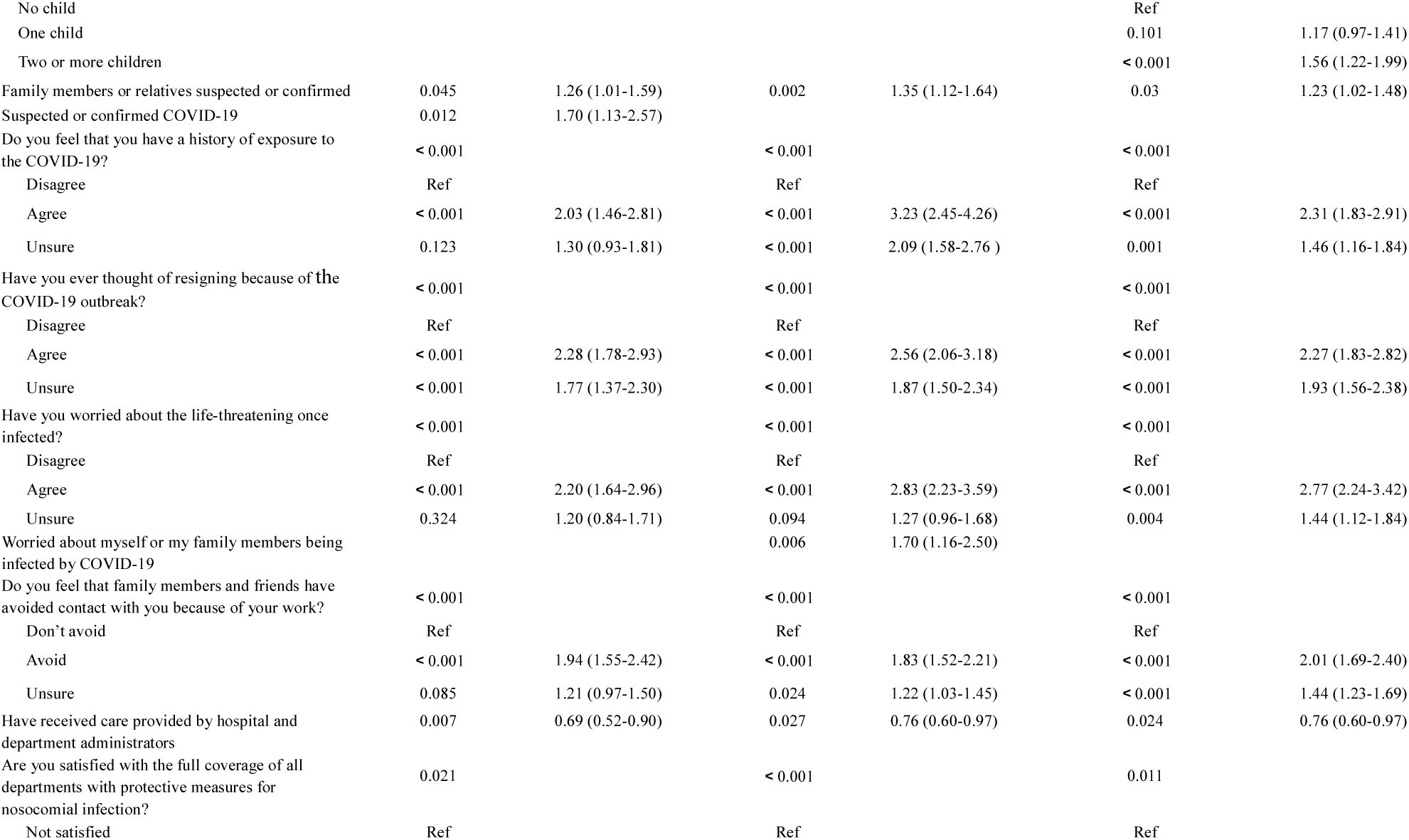
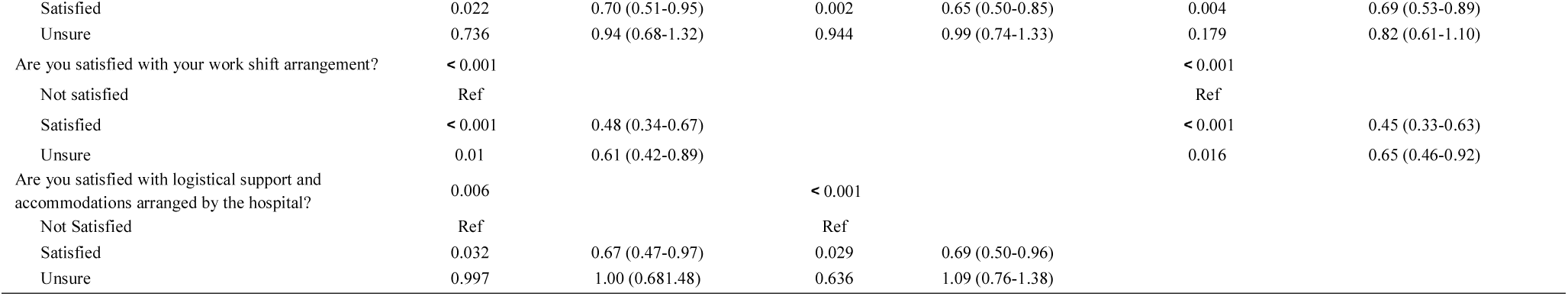
Factors associated with the depression, anxiety and psychological stress in Wuhan HWs dealing with the COVID-19

Four common risk factors of the perceptions of the threat of COVID-19 in three outcomes were also revealed (all P values < 0.001, Table 3): feel of exposure; resigning thought; feel of life-threatening; feel of family members and friends’ avoidance.

Two common protective factors of psychological protective measures in the three outcomes were presented (all P values<0·05, Table 3). Take stress outcome for example: care provided by hospital and department administrators (odds ratio [OR], 0.76; P=0.024) and full coverage of all departments with protective measures for nosocomial infection (OR, 0.69; P=0.004) were protective factors.

Moreover, reasonable work shift arrangement (OR, 0.69; P<0·001), sufficient logistical support and comfortable accommodations provided by hospital (OR, 0.67; P=0·03) were another two protective factors of depression symptoms. Drinking history (HR, 1.63; P=0.003) and suspected or confirmed COVID-19 (HR, 1.70; P=0.012) were two risk factors of depression outcomes.

In subgroup analysis, the influencing factors of the HWs who were confirmed or suspected patients (Group 1, n=158) were: resigning thought; feel of life-threatening; sufficient logistical support and comfortable accommodations arranged by hospital (all P values<0.01). The influencing factors of the HWs who had confirmed or suspected person in family members (Group 2, n=693 after excluding 70 HWs who were crossed with Group 1) were: age; past medical history; feel of exposure; resigning thought; feel of family members and friends’ avoidance; sufficient logistical support and comfortable accommodations arranged by hospital (all P values<0.05). The influencing factors of the others (Group 3, n=4211) were: age, professional title level, years of working, past medical history and other 7 variables of perceptions and psychological protective measures (all P values<0.05). The results of subgroup analysis were presented in Supplementary Appendix Tables S3, S6 and S9.

Living with family members (HR, 1.18; P=0.04) and worried about myself or my family members being infected by COVID-19 (HR, 1.70; P=0.006) were two risk factor and exercise habit was a protective factor (OR, 0.71; P=0.001) of anxiety symptoms. Sufficient logistical support and comfortable accommodations arranged by hospital was also a protective factor (OR, 0.69; P=0.03). There were 3, 7 and 13 influencing factors associated with anxiety symptoms in Group 1, 2 and 3 respectively, which were also presented in Supplementary Appendix Tables S3, S6 and S9.

Similarly, these factors were also found associated with psychological stress outcome: master degree or higher level; occupation; current work place; parent status; reasonable work shift arrangement (all P values<0.05). There were 2, 8 and 12 influencing factors with psychological stress in Group 1, 2 and 3 respectively, which were also presented in Supplementary Appendix Tables S3, S6 and S9.

### NOMOGRAMS CONSTRUCTION AND INTERNAL VALIDATION

We created three nomograms to predict the risk of three outcomes (Figure 1). Each variable is projected upward to the value of the Points line to get the score of each parameter. The total score was calculated by adding each score and located on the Total Points line. The risk was obtained according to the total score by drawing vertical lines to the Risk line. The C-statistics for depression, anxiety and psychological stress nomograms was 0.779 (95% CI, 0.775-0.783), 0.765 (95% CI, 0.761-0.769) and 0.766 (95% CI, 0.762-0.770), respectively. The calibration curves are plotted in Figure 2. Dots on the plots are very close to the 45° diagonal line, which suggests that the three nomograms were well-calibrated.

**Figure 1.**
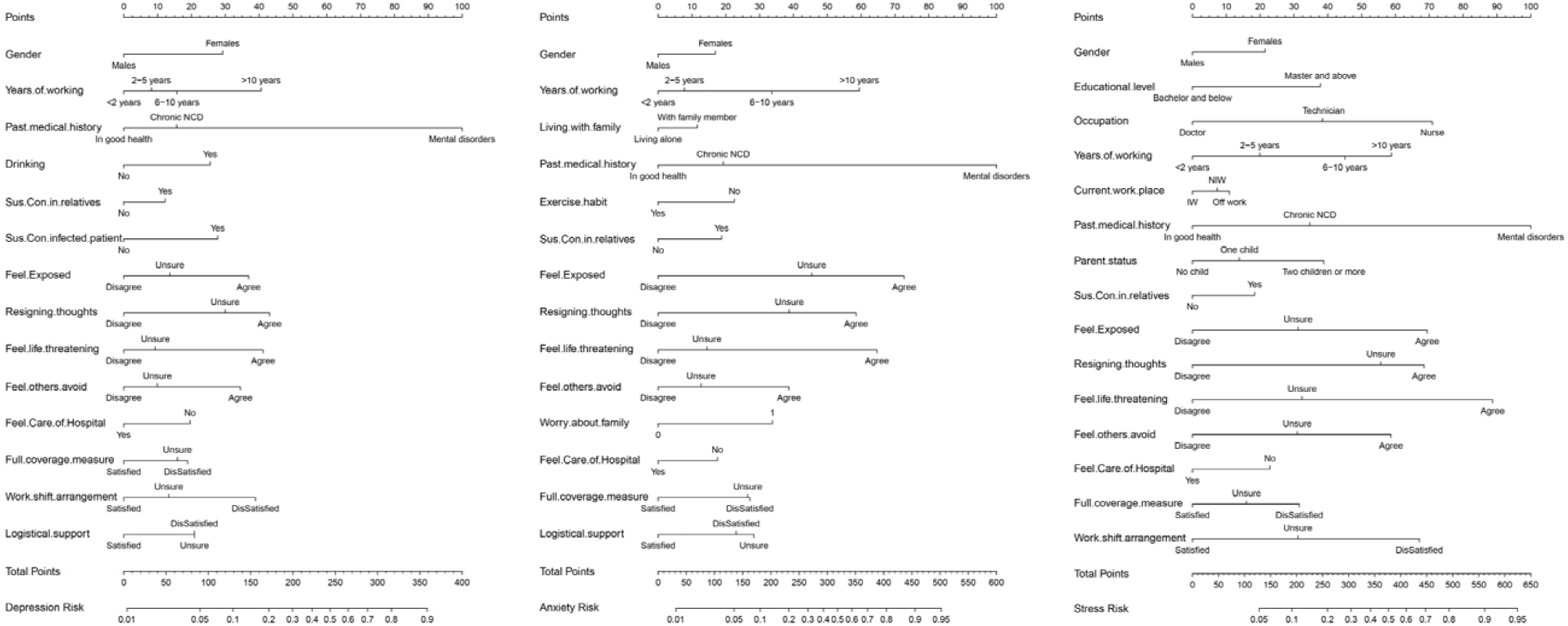
Nomograms of Depression, Anxiety and Stress

**Figure 2.**
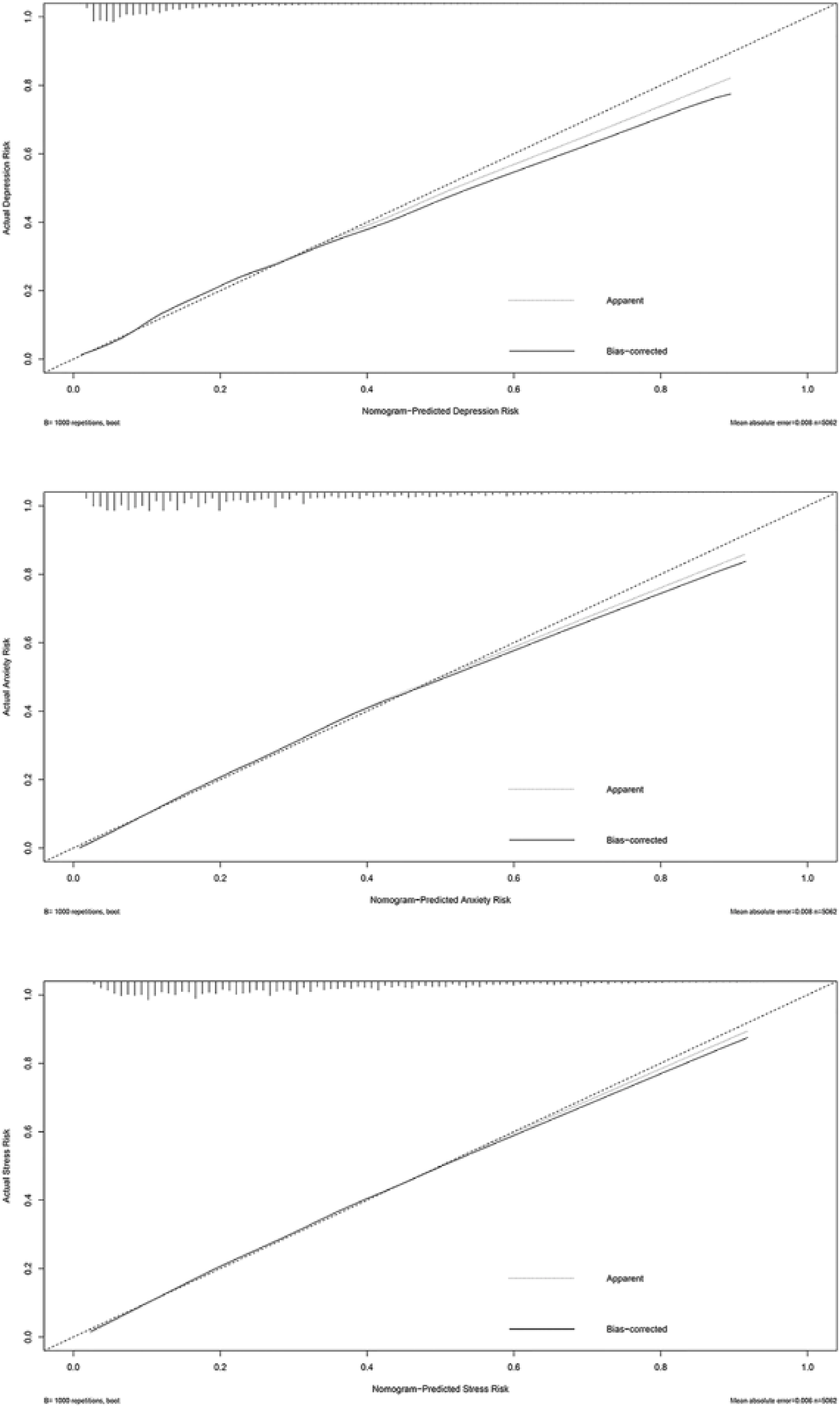
Calibration curves for Depression, Anxiety and Stress Nomograms

## DISCUSSION

Wuhan is the epicenter of COVID-19 epidemic with the most numerous and concentrated infected cases. The rising numbers of cases and deaths, coupled with the unprecedented lockdown of Wuhan, might create and spread public fear, panic, and distress. HWs at Tongji Hospital, officially designated as “the specific hospital for the treatment of severe patients with COVID-19 in Wuhan”, might be facing a serious psychological challenge.

Our single-center, cross-sectional survey showed that 1509 (29.8 percent), 681 (13.5 percent) and 1218 (24.1 percent) HWs reported stress, depression, and anxiety symptoms. A study during Taiwan’s SARS outbreak, which included 1257 HWs in 2003, showed 77.4 percent of HWs reported anxiety and worrying, and 74.2 percent of HWs reported depression. As well, a study involving 652 frontline medical staff showed that 68 percent of participants reported severe levels of job-related stress, and 57.0 percent were found to have experienced psychological distress during the Hong Kong SARS outbreak in 2003. Compared to the results of previous studies in SARS-affected hospitals^15-17^, the morbidity of depression and anxiety of HWs in this study was relatively lower, which may be related to the different measurements used in these studies and psychological protective measures implemented by Tongji Hospital’s administrators responding to COVID-19 in the early stage.

The management of Tongji Hospital was initially alert to the mental health problems that HWs might encounter, and therefore implemented a series of psychological protective measures. Hospital-based, department-based, and ward-based care provided by hospital and department administrators, and full coverage of all departments with protective measures for nosocomial infection were the common independent protective factors for acute stress, depression and anxiety. Although HWs are currently facing inconvenient transportation and shortage of safeguards, in this study, 83.6 percent of HWs were satisfied with sufficient logistical support and comfortable accommodations provided by Tongji Hospital, 79.5 percent were satisfied with the full coverage of all departments with protective measures for nosocomial infection, and 91.0 percent of HWs had received hospital-based, department-based, and ward-based care provided by hospital administrators and department leaders. Furthermore, 83.0 percent of HWs were satisfied with reasonable work shift arrangement, which was an independent protective factor for acute stress and depression. The hospital’s psychiatric team had also tried to support staff with relaxation techniques through online WeChat Balint group. However, only 5.0 percent of HWs joined in WeChat Balint group, suggesting that more psychosocial interventions and follow-up programs especially for the susceptible population need to be developed and promoted to reduce the perception of threats of COVID-19 among HWs.

Among the sociodemographic characteristics, the common risk factors for acute stress, depression, and anxiety symptoms were female, with history of mental disorders, history of physiological chronic non-communicable diseases, and family members or relatives suspected or confirmed COVID-19, and years of working >10 years. Gender factors and concomitant chronic non-communicable diseases have already been widely discussed and a considerable number of studies have suggested that females and people with concomitant chronic non-communicable diseases have higher risk of depression, anxiety and psychological stress^18-20^. Meanwhile, our study showed the risk of anxiety, depression, and acute stress tended to increase with increasing years of work, probably because most HWs with shorter working years were single (70.0 percent of HWs with years of working <2 years are single), with less occupational exhaustion and family responsibilities.

During the SARS period, 21.0 percent of the infected people were HWs^21^, while the proportion of infected HWs in Wuhan is approximately 4 percent^22^. In this study, 158 (3.1 percent) HWs were diagnosed or suspected COVID-19, and 70 of them (44.3 percent) had family members and relatives infected. In addition, 693 (15.1 percent) HWs were not infected themselves, but their relatives and family members were diagnosed or suspected of COVID-19. HWs had a significantly increased risk of depression once infected with the COVID-19 virus, and increased risks of anxiety, depression, and stress once their family members or relatives were infected with the COVID-19 virus. The results of the subgroup analyses provided more details and indicated that we should employ stratified psychological intervention strategies according to different risk levels of HWs. We should keep track of the mental status of those HWs who were confirmed or suspected patients themselves and who had confirmed or suspected patients in their families.

Consistent with the results of previous studies in SARS-affected and H1N1 influenza-affected hospitals, our study showed that nurses and medical technicians presented higher rates of psychological stress than doctors, probably because they have more and closer contact with patients. Meanwhile, in line with the results of previous studies in the SARS isolation unit, we also found that HWs in isolation wards have a more pronounced risk of stress^23^. Those who had two or more children showed a higher risk of stress, probably owing to their heavier family responsibilities. To make matters worse, the majority of HWs are female (85.0 percent in this study), since a large number of studies have confirmed the plight of working women^18^. Priority consideration should be given to female HWs. In our study, the exercise habit was associated with a lower risk of anxiety symptoms, suggesting that physical activity helps alleviate psychological impact caused by catastrophic events. Nevertheless, only 857 (16.9 percent) participants had exercise habits, indicating that the importance of exercise should be highlighted among HWs in China.

Finally, we developed three effective, easy-to-use clinical screening tools to identify high-risk individuals of acute stress, depression, and anxiety among HWs. Further studies are needed to improve and validate these models. Tongji Hospital’s experience suggests that psychological impact on HWs can be alleviated by timely implementation of psychological protective measures.

This work was supported by the National Key Research and Development Program of China (grant number 2017YFC1310000).

We acknowledge all participants of this project and investigators for collecting data.

## Data Availability

The de-identified database used in the current study are available from the corresponding authors at on reasonable request.

